# Metabolic changes induced by pharmacological castration of young, healthy men: a study of the plasma metabolome

**DOI:** 10.1101/2022.03.08.22271577

**Authors:** Jéssica de Siqueira Guedes, Indira Pla, K. Barbara Sahlin, Gustavo Monnerat, Roger Appelqvist, György Marko-Varga, Aleksander Giwercman, Gilberto Barbosa Domont, Aniel Sanchez, Fábio César Sousa Nogueira, Johan Malm

## Abstract

Testosterone is a hormone that plays a key role in carbohydrate, fat, and protein metabolism. Testosterone deficiency is associated with multiple comorbidities, *e.g*., metabolic syndrome and type 2 diabetes. Despite its importance in many metabolic pathways, the mechanisms by which it controls metabolism are not fully understood. The present study investigated the short-term metabolic changes of pharmacologically induced castration and testosterone supplementation in healthy young males. Thirty subjects were submitted to testosterone depletion (TD) followed by testosterone supplementation (TS). Plasma samples were collected three times corresponding to basal, low, and restored testosterone levels. An untargeted metabolomics study was performed by liquid chromatography-high resolution mass spectrometry (UHPLC-HRMS) to monitor the metabolic changes induced by the altered hormone levels. Our results demonstrated that TD is associated with major metabolic changes partially restored by TS. Carnitine and amino acid metabolism were the metabolic pathways most impacted by variations in testosterone. Furthermore, our results also indicate that LH and FSH might strongly alter the plasma levels of indoles and lipids, especially glycerophospholipids and sphingolipids. Our results demonstrate major metabolic changes induced by low testosterone that may be important for understanding the mechanisms behind the association of testosterone deficiency and its comorbidities.

## Introduction

Testosterone is the main androgen in men and plays diverse and essential roles in the human body, not only in reproduction but also in the modulation of carbohydrate, protein, and lipid metabolism ^1^. Testosterone has been associated with glucose uptake stimulation and insulin-regulated glucose transporter 4 (GLUT4) translocation in skeletal muscle cells, cardiomyocytes, and adipocytes ^2–4^. Other effects include an increment in lipolysis and lipid oxidation, which impacts the regulation of glucose metabolism and insulin function ^5^, and controlling the muscle mass to an anabolic state, increasing protein synthesis and decreasing protein breakdown ^6^.

Low testosterone levels have been associated with adverse metabolic profiles linked to hypogonadism, cardiovascular disease, metabolic syndrome, and diabetes ^7–10^. Moreover, patients with reduced androgen production and/or bioavailability often have erectile dysfunction (ED), increased fat mass index, decreased muscle strength, cognitive impairment, and mood disorders ^11,12^. Studies on testosterone replacement therapy (TRT) have demonstrated improved quality of life in men with hypogonadism, reduced symptoms such as ED and fat mass index, and increased lean body mass and sexual desire^13,14^. Furthermore, TRT improved insulin resistance and glycemic control in men with testosterone deficiency ^15,16^.

Despite the multifaceted functions of testosterone in reproduction and metabolism and the diverse comorbidities associated with low testosterone, the molecular changes induced by testosterone deficiency have not been fully characterized. A better understanding of these events would make possible not only a comprehension of the pathobiology but also identification of new biomarkers, improved diagnostic strategies and methods, and monitoring of treatment.

In this context, metabolomics has been suggested as a valuable strategy for investigation of disease and response to testosterone supplementation ^17^. MS-based untargeted metabolomics is a powerful tool for metabolic and endocrine screening and has the potential to identify new biomarkers suitable for implementation in patient care ^18,19^.

Our group previously designed a short-term human model where thirty healthy young males were submitted to a testosterone depletion followed by testosterone supplementation. Blood samples corresponding to basal, low, and restored testosterone levels were collected ^20^. Proteomics analysis of this controlled cohort revealed biological processes driven by gonadotropin and testosterone ^21^ and indicated proteins and amino acids associated with testosterone fluctuations ^22,23^. However, a deep metabolomics study associated with testosterone and gonadotropins changes was not performed.

This study aims to describe a deep plasma metabolic profile in healthy young men with pharmacologically induced alterations in testosterone and gonadotropins levels. Plasma samples from the human model were analyzed by MS-based untargeted metabolomics. Our results highlight for the first time the complex metabolic changes induced by androgen depletion in healthy men and the short-term effect of testosterone recovery in this cohort. Several metabolites were impacted, especially those related to carnitine and amino acid metabolism. On the other hand, many metabolites associated with indoles and lipid metabolism appear to be affected by gonadotropin, *i.e*., LH and FSH, rather than testosterone levels.

## Results

Our untargeted metabolomics analysis (UHPLC-HRMS) on plasma from thirty healthy young men submitted to gonadotropin and testosterone deficiency (TD) followed by testosterone supplementation (TS) identified 706 metabolites that were altered as a result of pharmacologically induced hormonal changes. The samples were classified into three groups: Basal, Low, and Restored testosterone levels (Figure 1), and the metabolites were categorized according to their chemical class (Figure 2A).

**Figure 1.**
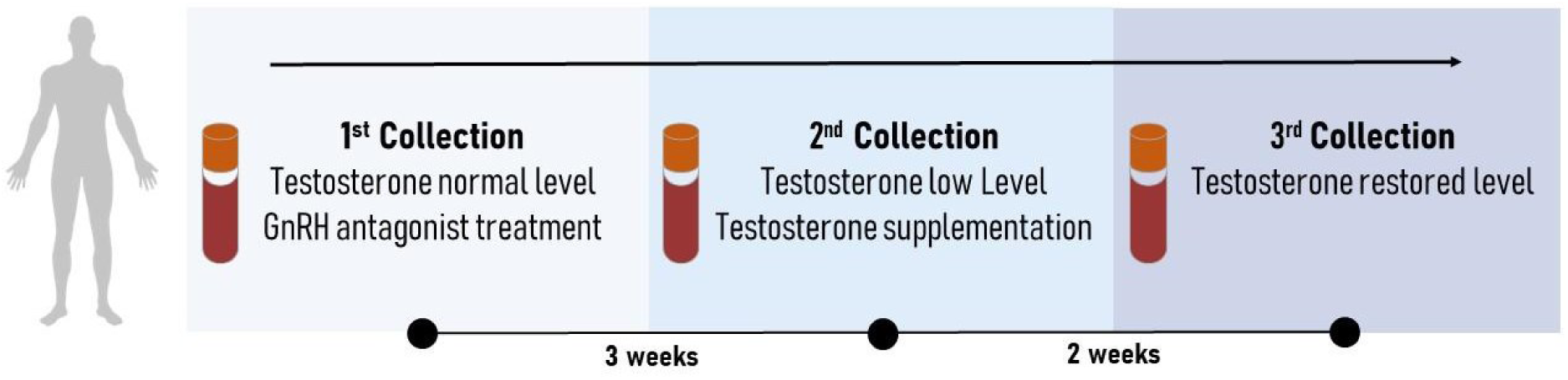
Schematic representation of the designed human model. Blood samples were collected three times. The first collection was done before any pharmacological treatment and represents testosterone and gonadotropins at baseline levels. During the same visit, after blood collection, all subjects received a subcutaneous injection of gonadotropin-releasing hormone antagonist (GnRHa, 240 mg, Degarelix®, Ferring Pharmaceuticals, Sweden). Three weeks later, subjects’ blood samples were collected again, corresponding to low testosterone and gonadotropin levels. All participants then received an intramuscular injection of testosterone undecanoate (1000mg, Nebido®, Bayer Pharmaceuticals, Germany). After two weeks, the last blood samples were collected, corresponding to testosterone restored samples and low gonadotropin levels.

**Figure 2.**
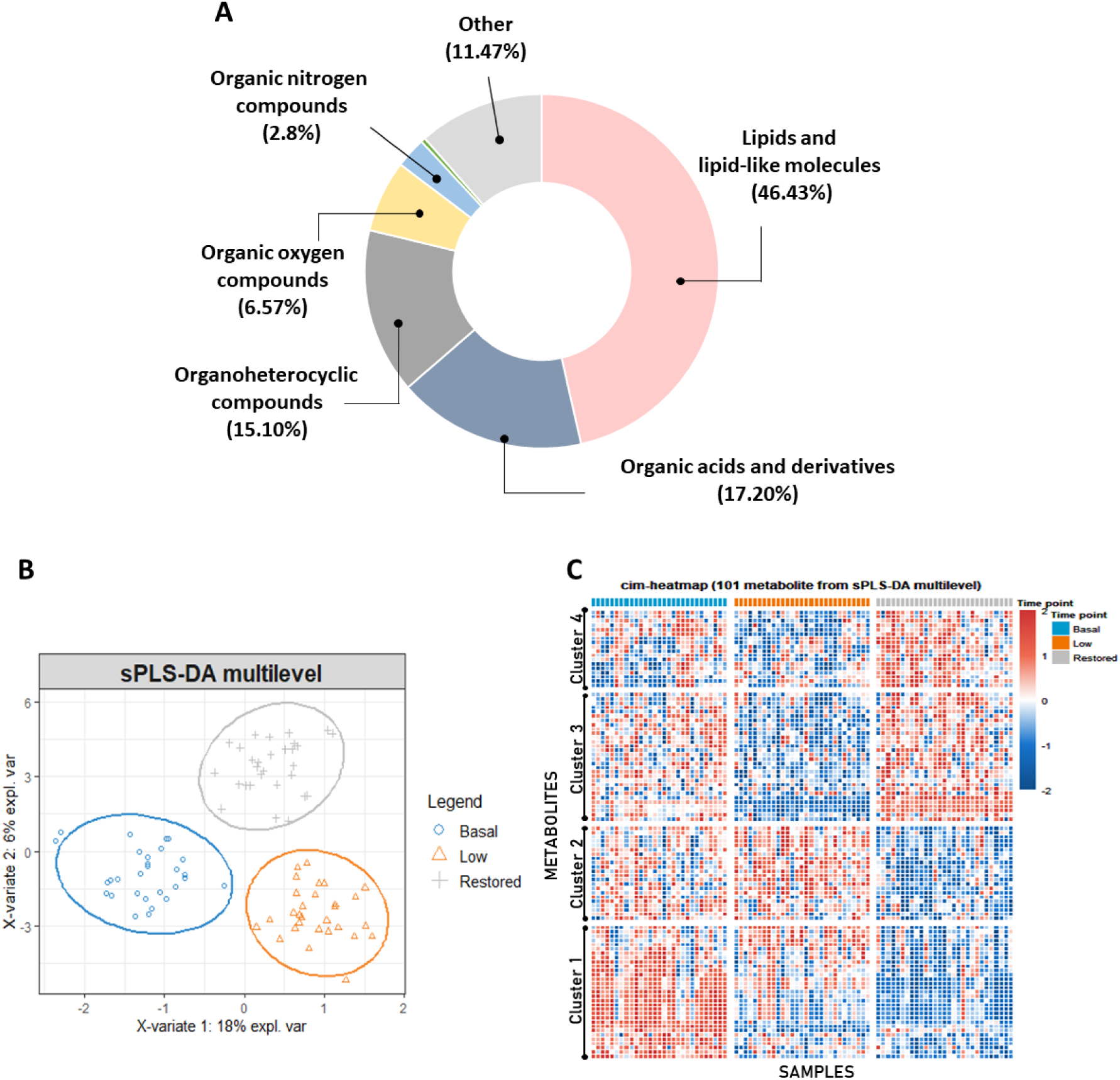
Untargeted metabolomics analysis. A) Chemical class of compounds identified by untargeted metabolomics approach based on UHPLC-HRMS. B) Multilevel multivariate approach: sPLS-DA 2D score plot. The blue, orange, and gray regions represent the 95% confidence area of each group. The blue circles represent the Basal testosterone group, the orange triangles symbolize the Low testosterone group after chemical castration, and the gray crosses represent the Restored testosterone group after TS. C) hierarchical clustering and heatmap based on sPLS-DA data. Four major metabolites clusters were revealed. Most compounds present in clusters 2, 3 and 4 seem to be driven by testosterone since they recovered or tended to recover their levels after TS. Conversely, many metabolites present in cluster 1 appear to be more influenced by LH and FSH expression since their levels did not normalize after TS.

A Sparse Partial Least Squares Discriminant Analysis (sPLS-DA) discriminated the samples according to the sample collection time points, *i.e*., different testosterone levels (Basal, Low, and Restored) (Figure 2B), emphasizing the particular metabolic signature of each group. The hierarchical clustering analysis based on sPLS-DA data revealed four distinct clusters, highlighting some key metabolites (Figure 2C). Table 1 shows the fifty known compounds present in the Human Metabolome Database (HMDB) and/or in the Kyoto Encyclopedia of Genes and Genomes (KEGG) that were responsible for the cluster separation, among them only six were non-significant (ANOVA paired analysis, *q*-value > 0.05). Other molecules were not included. Additional information regarding the compounds identified by sPLS-DA, such as normalized average area value and Log2 fold-change, are present in the supplementary data (Table S1).

**Table 1.**
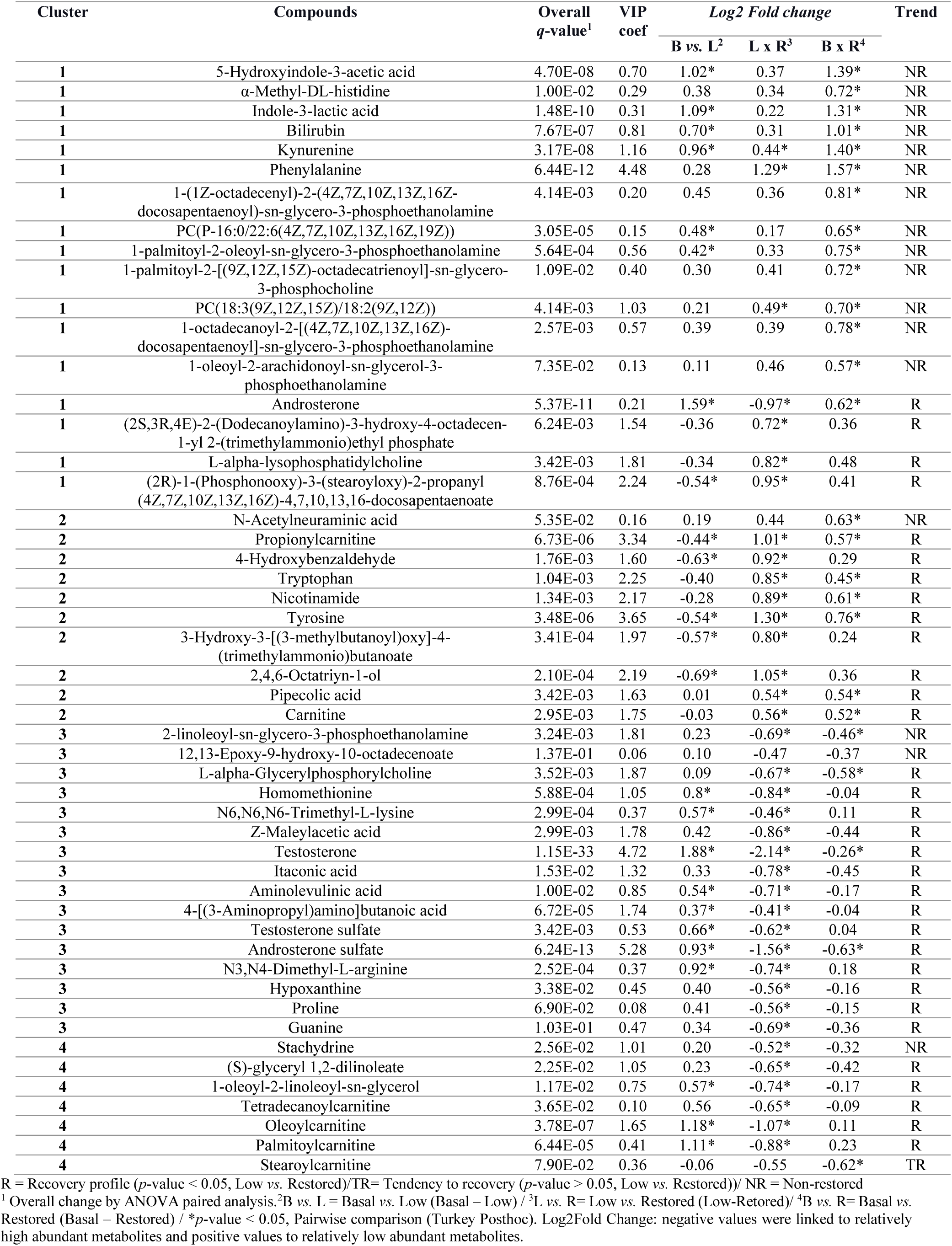
Metabolites identified by sPLS-DA analysis

| Cluster | Compounds | Overall<br>*q*-value<sup>1</sup> | VIP<br>coef | Log2 Fold change |  |  | Trend |
| --- | --- | --- | --- | --- | --- | --- | --- |
|  |  |  |  | B vs. L<sup>2</sup> | L x R<sup>3</sup> | B x R<sup>4</sup> |  |
| 1 | 5-Hydroxyindole-3-acetic acid | 4.70E-08 | 0.70 | 1.02* | 0.37 | 1.39* | NR |
| 1 | α-Methyl-DL-histidine | 1.00E-02 | 0.29 | 0.38 | 0.34 | 0.72* | NR |
| 1 | Indole-3-lactic acid | 1.48E-10 | 0.31 | 1.09* | 0.22 | 1.31* | NR |
| 1 | Bilirubin | 7.67E-07 | 0.81 | 0.70* | 0.31 | 1.01* | NR |
| 1 | Kynurenine | 3.17E-08 | 1.16 | 0.96* | 0.44* | 1.40* | NR |
| 1 | Phenylalanine | 6.44E-12 | 4.48 | 0.28 | 1.29* | 1.57* | NR |
| 1 | 1-(1Z-octadecenyl)-2-(4Z,7Z,10Z,13Z,16Z-docosapentaenyl)-sn-glycero-3-phosphoethanolamine | 4.14E-03 | 0.20 | 0.45 | 0.36 | 0.81* | NR |
| 1 | PC(P-16:0/22:6(4Z,7Z,10Z,13Z,16Z,19Z)) | 3.05E-05 | 0.15 | 0.48* | 0.17 | 0.65* | NR |
| 1 | 1-palmitoyl-2-oleoyl-sn-glycero-3-phosphoethanolamine | 5.64E-04 | 0.56 | 0.42* | 0.33 | 0.75* | NR |
| 1 | 1-palmitoyl-2-[(9Z,12Z,15Z)-octadecatrienoyl]-sn-glycero-3-phosphocholine | 1.09E-02 | 0.40 | 0.30 | 0.41 | 0.72* | NR |
| 1 | PC(18:3(9Z,12Z,15Z)/18:2(9Z,12Z)) | 4.14E-03 | 1.03 | 0.21 | 0.49* | 0.70* | NR |
| 1 | 1-octadecanoyl-2-[(4Z,7Z,10Z,13Z,16Z)-docosapentaenyl]-sn-glycero-3-phosphoethanolamine | 2.57E-03 | 0.57 | 0.39 | 0.39 | 0.78* | NR |
| 1 | 1-oleoyl-2-arachidonoyl-sn-glycerol-3-phosphoethanolamine | 7.35E-02 | 0.13 | 0.11 | 0.46 | 0.57* | NR |
| 1 | Androsterone | 5.37E-11 | 0.21 | 1.59* | -0.97* | 0.62* | R |
| 1 | (2S,3R,4E)-2-(Dodecanoylamino)-3-hydroxy-4-octadecen-1-yl 2-(trimethylammonio)ethyl phosphate | 6.24E-03 | 1.54 | -0.36 | 0.72* | 0.36 | R |
| 1 | L-alpha-lysophosphatidylcholine | 3.42E-03 | 1.81 | -0.34 | 0.82* | 0.48 | R |
| 1 | (2R)-1-(Phosphonooxy)-3-(stearoyloxy)-2-propanyl (4Z,7Z,10Z,13Z,16Z)-4,7,10,13,16-docosapentaenoate | 8.76E-04 | 2.24 | -0.54* | 0.95* | 0.41 | R |
| 2 | N-Acetylneuraminic acid | 5.35E-02 | 0.16 | 0.19 | 0.44 | 0.63* | NR |
| 2 | Propionylcarnitine | 6.73E-06 | 3.34 | -0.44* | 1.01* | 0.57* | R |
| 2 | 4-Hydroxybenzaldehyde | 1.76E-03 | 1.60 | -0.63* | 0.92* | 0.29 | R |
| 2 | Tryptophan | 1.04E-03 | 2.25 | -0.40 | 0.85* | 0.45* | R |
| 2 | Nicotinamide | 1.34E-03 | 2.17 | -0.28 | 0.89* | 0.61* | R |
| 2 | Tyrosine | 3.48E-06 | 3.65 | -0.54* | 1.30* | 0.76* | R |
| 2 | 3-Hydroxy-3-[(3-methylbutanoyl)oxy]-4-(trimethylammonio)butanoate | 3.41E-04 | 1.97 | -0.57* | 0.80* | 0.24 | R |
| 2 | 2,4,6-Octatriyn-1-ol | 2.10E-04 | 2.19 | -0.69* | 1.05* | 0.36 | R |
| 2 | Pipecolic acid | 3.42E-03 | 1.63 | 0.01 | 0.54* | 0.54* | R |
| 2 | Carnitine | 2.95E-03 | 1.75 | -0.03 | 0.56* | 0.52* | R |
| 3 | 2-linoleoyl-sn-glycero-3-phosphoethanolamine | 3.24E-03 | 1.81 | 0.23 | -0.69* | -0.46* | NR |
| 3 | 12,13-Epoxy-9-hydroxy-10-octadecenoate | 1.37E-01 | 0.06 | 0.10 | -0.47 | -0.37 | NR |
| 3 | L-alpha-Glycerolphosphorylcholine | 3.52E-03 | 1.87 | 0.09 | -0.67* | -0.58* | R |
| 3 | Homomethionine | 5.88E-04 | 1.05 | 0.8* | -0.84* | -0.04 | R |
| 3 | N6,N6,N6-Trimethyl-L-lysine | 2.99E-04 | 0.37 | 0.57* | -0.46* | 0.11 | R |
| 3 | Z-Maleylacetic acid | 2.99E-03 | 1.78 | 0.42 | -0.86* | -0.44 | R |
| 3 | Testosterone | 1.15E-33 | 4.72 | 1.88* | -2.14* | -0.26* | R |
| 3 | Itaconic acid | 1.53E-02 | 1.32 | 0.33 | -0.78* | -0.45 | R |
| 3 | Aminolevulinic acid | 1.00E-02 | 0.85 | 0.54* | -0.71* | -0.17 | R |
| 3 | 4-[(3-Aminopropyl)amino]butanoic acid | 6.72E-05 | 1.74 | 0.37* | -0.41* | -0.04 | R |
| 3 | Testosterone sulfate | 3.42E-03 | 0.53 | 0.66* | -0.62* | 0.04 | R |
| 3 | Androsterone sulfate | 6.24E-13 | 5.28 | 0.93* | -1.56* | -0.63* | R |
| 3 | N3,N4-Dimethyl-L-arginine | 2.52E-04 | 0.37 | 0.92* | -0.74* | 0.18 | R |
| 3 | Hypoxanthine | 3.38E-02 | 0.45 | 0.40 | -0.56* | -0.16 | R |
| 3 | Proline | 6.90E-02 | 0.08 | 0.41 | -0.56* | -0.15 | R |
| 3 | Guanine | 1.03E-01 | 0.47 | 0.34 | -0.69* | -0.36 | R |
| 4 | Stachydrine | 2.56E-02 | 1.01 | 0.20 | -0.52* | -0.32 | NR |
| 4 | (S)-glyceryl 1,2-dilinoate | 2.25E-02 | 1.05 | 0.23 | -0.65* | -0.42 | R |
| 4 | 1-oleoyl-2-linoleoyl-sn-glycerol | 1.17E-02 | 0.75 | 0.57* | -0.74* | -0.17 | R |
| 4 | Tetradecanoylcarnitine | 3.65E-02 | 0.10 | 0.56 | -0.65* | -0.09 | R |
| 4 | Oleoylcarnitine | 3.78E-07 | 1.65 | 1.18* | -1.07* | 0.11 | R |
| 4 | Palmitoylcarnitine | 6.44E-05 | 0.41 | 1.11* | -0.88* | 0.23 | R |
| 4 | Stearoylcarnitine | 7.90E-02 | 0.36 | -0.06 | -0.55 | -0.62* | TR |

R = Recovery profile ( $p$ -value < 0.05, Low vs. Restored)/TR= Tendency to recovery ( $p$ -value > 0.05, Low vs. Restored)/ NR = Non-restored
<sup>1</sup> Overall change by ANOVA paired analysis. <sup>2</sup>B vs. L = Basal vs. Low (Basal – Low) / <sup>3</sup>L vs. R= Low vs. Restored (Low-Retored)/ <sup>4</sup>B vs. R= Basal vs. Restored (Basal – Restored) / \* $p$ -value < 0.05, Pairwise comparison (Turkey Posthoc). Log2Fold Change: negative values were linked to relatively high abundant metabolites and positive values to relatively low abundant metabolites.

The hierarchical clustering analysis demonstrates some metabolites reverted to basal levels or presented a tendency to recover after testosterone replacement, indicating that these compounds could potentially reflect the plasma testosterone level. In this respect, most compounds present in clusters 2, 3, and 4 seem to be driven by testosterone (Figure 2C and Table 1), among them tyrosine, tryptophan, oleoylcarnitine, palmitoylcarnitine, and testosterone metabolites (ANOVA paired analysis, *q*-value ≤ 0.05). Moreover, the biological roles connected with these metabolites present in clusters 2, 3, and 4 highlighted the importance of carnitine, beta-oxidation of fatty acids, and amino acid metabolism (*p*-value > 0.05) as the top metabolic pathways altered by testosterone levels (Figure 3). Moreover, other molecules (cluster 1) appear to be influenced by gonadotrophins, *i.e*., LH and FSH, since their levels did not restore after TS (Figure 2C, Table 1). This scenario includes altered indole compounds and different glycerophospholipids (ANOVA paired analysis, (*q*-value ≤ 0.05).

**Figure 3.**
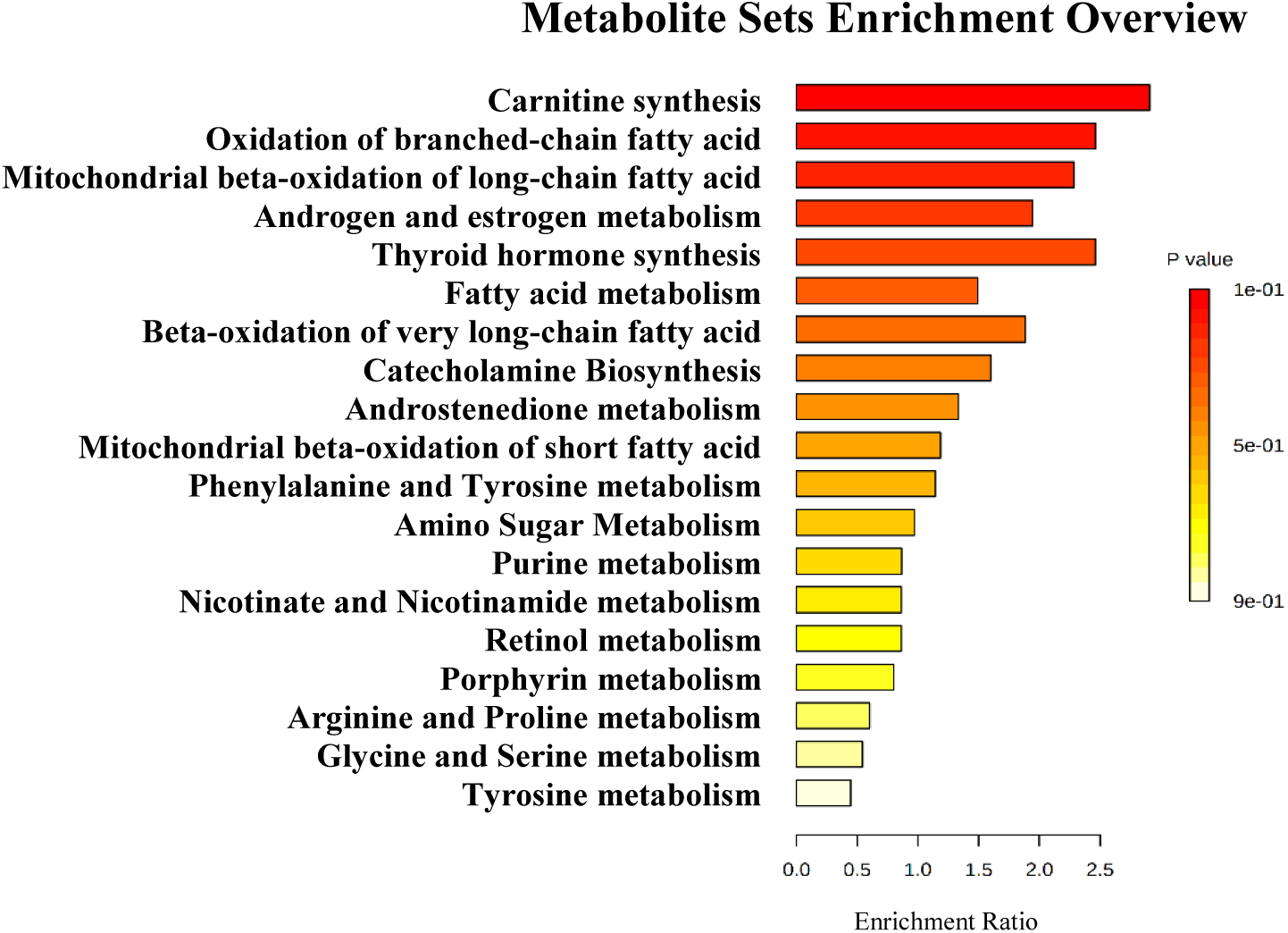
Pathway analysis of markers metabolites of testosterone levels identified by sPLS-DA analysis. Metabolic Set Enrichment Analysis (MSEA) results based on search in The Small Molecule Pathway Database (SMPDB) library. Bar graphs illustrating the enrichment overview of the top metabolic pathways of metabolites present in clusters 2, 3, and 4 with recovery or tendency to recover the respective abundance profiles. Color intensity (yellow to red) indicates their increasing statistical significance.

Additionally, we carried out an ANOVA paired analysis to deeply examine which compounds were most impacted by the different plasma testosterone levels. A total number of 368 compounds with a statistical difference (*q-*value ≤ 0.05, ANOVA paired) were identified, among them 311 were altered by low levels of testosterone and gonadotropins (pairwise comparison, Basal *vs*. Low), 83 molecules normalized after testosterone supplementation (*p*-value ≤ 0.05, Low *vs*. Restored) and 98 demonstrated tendencies to recovery based on their median values (not statistically significant, Low *vs*. Restored).

The metabolites present in the Human Metabolome Database (HMDB) that appear to be regulated by testosterone are listed in the Table 2 (recovery or tendency to recovery profile). Compounds that appear to be driven by gonadotropins (metabolites non-restored) are listed in Table 3. Information about the chemical class or subclass and suggested biological function of respective metabolites is included. Metabolites present in Table 1 were not included. All molecules, including non-significant compounds are present in the supplementary data (Table S2 and S3).

**Table 2.**
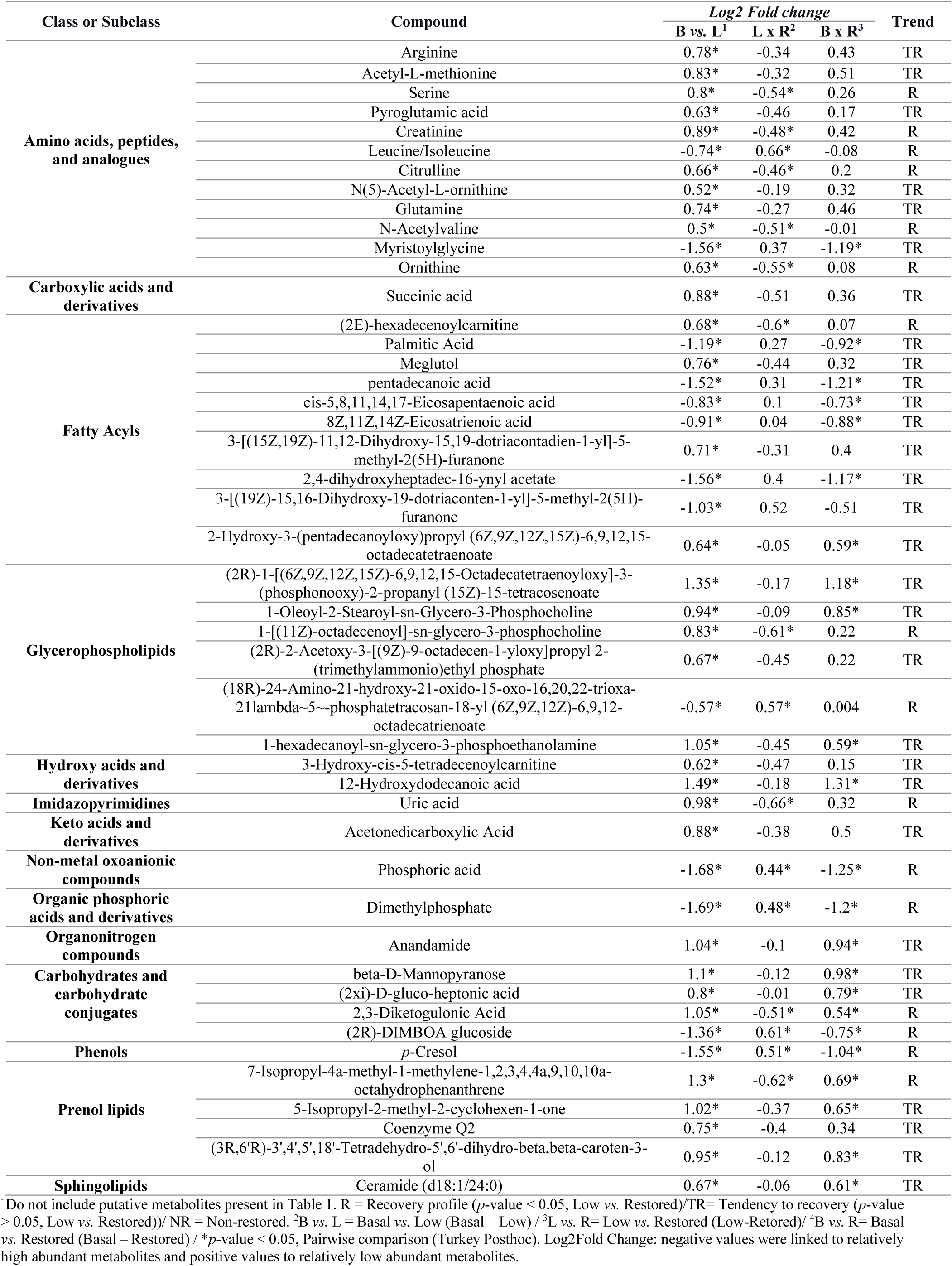
Metabolites associated with testosterone level^ǂ^

**Table 3.**
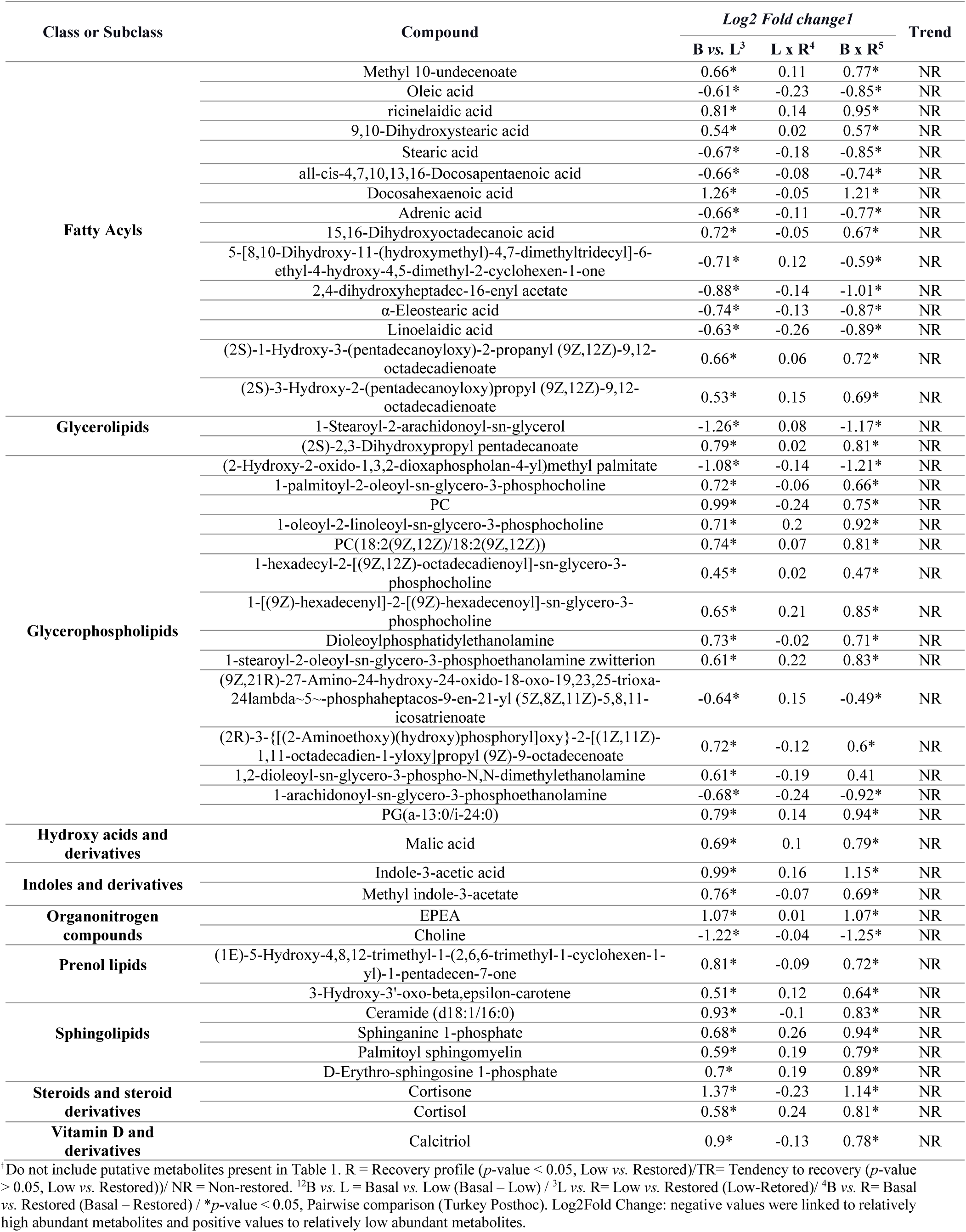
Metabolites associated with gonadotropin level^ǂ^

### Markers compounds of testosterone and gonadotropin levels

The untargeted metabolomics analysis revealed metabolites that vary in concentration in parallel with testosterone. Acylcarnitines, many intermediate compounds of arginine metabolism, and common biomarkers of renal function, *i.e*., creatinine and uric acid, decrease after TD and normalize after TS. On the other hand, aromatic amino acids, *i.e*., tryptophan and tyrosine, seem to be driven by testosterone in a converse manner, increasing in concentration after androgen depletion and reverting to basal levels with TS (Figure 4 and Figure S2).In contrast, cortisol, a hydroxysteroid, did not recover after TS (Figure S1).

**Figure 4.**
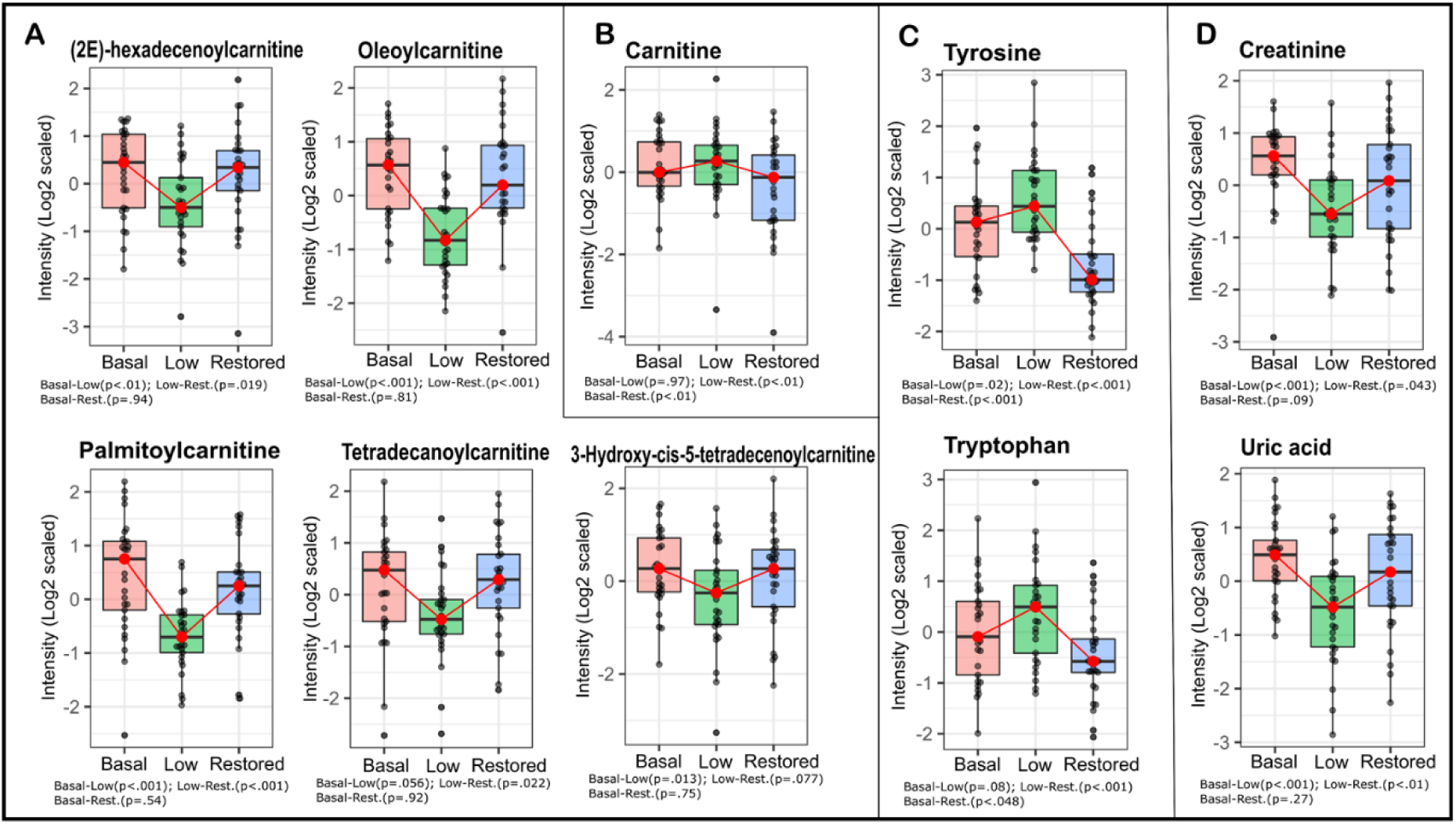
Markers of testosterone levels. Levels of some metabolites after TD and TS. A) Some acylcarnitines: (2E)-hexadecenoylcarnitine^*,#^, Oleoylcarnitine^*,#^, Palmitoylcarnitine^*,#^, 3-Hydroxy-cis-5-tetradecenoylcarnitine^*^, and Tetradecanoylcarnitine^*,#^. B) Carnitine^#,ǂ^. C) Aromatic amino acids: Tyrosine^*,#,ǂ^, and Tryptophan^#,ǂ^. D) Biomarkers of kidney function: Creatinine^*,#^, and Uric acid^*,#^. Boxplot colors: Pink, Basal testosterone group; Green, Low testosterone group; Blue, Restored testosterone group. \**p*-value ≤ 0.05, Basal *vs*. Low, ^#^*p*-value ≤ 0.05, Low *vs*. Restored, ^ǂ^*p*-value ≤ 0.05, Basal *vs*. Restored.

In addition to evaluating short-term metabolic changes driven by testosterone, our study model may also allow for identifying metabolites that reflect LH and FSH levels. The results indicate that low LH and FSH might alter the concentration of plasma lipids, *e.g*., glycerophospholipids and sphingolipids, compounds associated with tryptophan and indole metabolism, and calcitriol. Our results showed that these metabolites decreased in parallel with the gonadotropins (Table 3).

## Discussion

To the best of our knowledge, this is the first study investigating the metabolomic changes in a cohort of young, healthy men with pharmacologically induced TD and subsequent TS. Testosterone deficiency has a complex association with hypogonadism, obesity, metabolic dysfunction, impaired glucose tolerance, type 2 diabetes mellitus, and sleep disorders ^24^. The design of our study model allows an evaluation of the early metabolic effects associated with variations in testosterone and gonadotropins levels without the influence of comorbidities and older age. These changes correspond to what was seen with the present human model, both amino acid and lipid metabolism was altered, and testosterone supplementation could restore 83 of 311 metabolites (additionally, 98 compounds demonstrated tendencies to recovery), indicating that some of the metabolic changes may be driven by a decrease in gonadotropin rather than testosterone levels. Insulin-like growth factors, tyrosine catabolism and the level of many other amino acids, glucose metabolism (glycolysis/gluconeogenesis) ^21–23^, as well as steroidal profile, mitochondrial metabolism, and renal biomarkers were impacted by testosterone or its derivatives. The amino acid changes indicate the importance of testosterone for protein turnover. Our results further suggest that LH and FSH strongly affect the levels of indoles and lipids, especially glycerophospholipids and sphingolipids.

Carnitine is a key metabolite in energy metabolism in most mammals where it supports energy metabolism by transporting long-chain fatty acids into mitochondria for energy production. Our metabolomic analysis revealed that acylcarnitines levels change in a testosterone-sensitive direction, decreasing after TD and being restored by TS (Table 2 and Figure 4A). This is in agreement with Fanelli and coworkers who showed that male patients with hypogonadism present lower levels of acylcarnitines ^25^, an alteration that was even more pronounced in insulin resistance patients, which appears to be involved with the metabolism of acetyl-CoA and production of precursor for gluconeogenesis ^26^, and also with our previous study in rats where it was shown that TS increases the acylcarnitine levels ^27^.

Additionally, testosterone has been associated with the promotion of lipid oxidation ^28,29^. Our results showed that the concentration of palmitoylcarnitine and palmitic acid, lipid oxidation-related molecules, were correlated with testosterone changes. Palmitoylcarnitine decreased upon testosterone decrease and increased after testosterone supplementation, whereas palmitic acid increased upon testosterone decrease and was partially restored after testosterone supplementation. Similar changes were seen in a previously published study, where palmitic acid oxidation increased in human myotubes after treatment with testosterone ^30^.

Also, several studies have pointed out that testosterone could impair renal function, promoting tubular epithelial apoptosis in a dose-dependent way ^31^ and reducing estimated glomerular filtration, which could elevate creatinine levels ^32^. Our study also indicates that testosterone could promote higher levels of kidney function markers (*i.e*., creatinine and uric acid) since subjects with basal and restored testosterone levels showed an increased abundance of these metabolites (Table 2 and Figure 4D). These findings are in agreement with a recent study where higher testosterone was associated with an increment in uric acid levels in adolescent boys ^33^, and a case report of hypogonadism where renal function decreased after testosterone replacement therapy, increasing creatinine levels ^34^. However, considering that creatinine is also involved in muscle metabolism, the positive correlation between testosterone and creatinine might reflect the increase in muscle mass and not an impairment in renal function^35^. This is especially important, taking into account that creatinine has been reported as a biased biomarker of acute changes in kidney function since its levels can vary widely with hydration status, age, sex, muscle mass, and metabolism ^36^. Similarly, increased uric acid levels might mirror mass muscle growth due to a rise in purines levels ^37^.

Our results concerning amino acids and protein turnover are challeging. The hierarchical clustering analysis based on sPLS-DA data highlighted that the abundance of some amino acids (*e.g*. tryptophan and tyrosine) correlate negatively with testosterone, low levels of testosterone were associated with increased tryptophan and tyrosine levels, supporting the suppressive effect of testosterone on protein breakdown (Table 1 and Figure 4C) ^38^. This is in contrast to arginine and metabolites of the arginine biosynthesis pathway, especially citrulline and ornithine, that decreased in concentration after testosterone depletion and were restored or showed a tendency to restore (based on median values) after testosterone supplementation. Considering that citrulline and arginine have been reported as functional nutrients able to enhance muscle protein synthesis ^39–41^, these findings might reflect the muscle protein anabolism mediated by testosterone.

The results concerning lipid metabolism regulation shows complex metabolic picture. The ANOVA paired analysis and the sPLS-DA data highlighted that testosterone alteration and gonadotropins depletion are linked to lipid metabolism. While some of the lipid metabolites increased after androgen deprivation, others decreased. The different patterns may be due to the different effects of testosterone and gonadotropins, testosterone appears to suppress fatty acids synthesis ^42^, whereas FSH promotes the biosynthesis of these metabolites by upregulation of genes involved in lipid biosynthesis ^43,44^. In addition, most sphingolipids changed parallel with the gonadotropins, reduced levels after testosterone depletion and no normalization after testosterone supplementation, thus suggesting that FSH and/or LH are important for lipid metabolism (Table 2 and Table 3). Interestingly, sphingolipids (*e.g*., ceramides, sphingomyelin, and sphingosine-1-phosphate) appear to influence steroidogenesis at different biological levels, including actions as second messengers in signaling cascades and regulation of gene and enzymes expression that controls steroidogenesis ^45,46^.

The level of one of the fat soluble vitamins, calcitriol (*i.e*., bioactive vitamin D) showed a tendency to decrease upon testosterone depletion, and did not increase after testosterone supplementation, possibly indicating that gonadotropins are involved in vitamin D metabolism, also supported by the fact that low levels of vitamin D have been correlated with secondary hypogonadism ^21,47^. Moreover, according to the The European Society of Endocrinology Clinical Guideline several hormonal alterations, among them testosterone deficiency, low values of LH/FSH ratios in men, and vitamin D deficiency ^48^ often occur together, indicating that a functional relationship may exist.

Lastly, some metabolites of tryptophan and indoles turnover decreased after testosterone depletion and were not restored by testosterone. Some of these indoles represent microbial metabolites from tryptophan gut metabolism ^49^ and the alteration may be the result of the altered steroid profile, several studies indicate that steroids influence the gut microbiome ^50,51^.

In summary, our study demonstrates the importance of testosterone and gonadotropins for the plasma metabolome in young, healthy men. Major metabolic pathways were impacted by testosterone depletion, changes that only in some cases could be reverted by testosterone supplementation indicating that not only testosterone but also gonadotropins are important for metabolic regulation. These findings may contribute to a better understanding of the mechanisms behind the association of testosterone deficiency and its comorbidities.

## Materials and Methods

We performed an untargeted metabolomics analysis based on UHPLC-HRMS to evaluate the short-term metabolic changes associated with testosterone depletion and its subsequent restoration in a healthy young men cohort. The efficiency of chemical castration and testosterone replacement was previously verified by measuring the levels of gonadotropins and sex hormones (Table 4) ^20^. Most subjects restored their testosterone levels or exhibited higher testosterone than at baseline after testosterone supplementation. As expected, LH and FSH were not restored after testosterone supplementation due to the GnRHa effect.

**Table 4.** Plasma concentration of gonadotropins and sex hormones in the three groups: Basal, Low and Restored testosterone levels

| Biomarker | Mean ± SD | | |
| --- | --- | --- | --- |
|  | Basal | Low | Restored |
| Testosterone (nmol/L)<sup>a,b,c</sup> | 19.97 ± 5.7 | 0.71 ± 0.3 | 37.39 ± 11.1 |
| Estradiol (pmol/L)<sup>a,b,c</sup> | 107.66 ± 18.4 | 91 ± 0 | 144.45 ± 49.6 |
| Luteinising hormone -LH (IU/L)<sup>a,c</sup> | 4.99 ± 1.6 | 0.14 ± 0.1 | 0.14 ± 0.1 |
| Follicle stimulating hormone - FSH (IU/L)<sup>a,c</sup> | 3.09 ± 1.7 | 0.15 ± 0.1 | 0.14 ± 0.1 |

This results were extracted from K.B Sahlin. et.al. <sup>22</sup>. Variables are expressed as mean ± standard deviation
<sup>a</sup> *p*-value < 0.05, Basal vs. Low
<sup>b</sup> *p*-value < 0.05, Low vs. Restored
<sup>c</sup> *p*-value < 0.05, Basal vs. Restored

### Subjects and Ethical considerations

Thirty healthy men of 23.9 years old (19-32 years) were recruited for this study. All subjects had a body mass index of 20-25 kg/m^2^ and were non-smokers or occasional smokers, without any chronic diseases, such as cancer, diabetes, liver, and heart disease. The selected participants did not use anabolic steroids or any regular medication. All experimental procedures were registered and approved by the Regional Ethical Review Board Lund (Approval number: DNR 2014/ 311) and all participants signed informed consent ^20^.

### Study design: Testosterone depletion and replacement

Thirty subjects were submitted to a testosterone depletion by the use of gonadotropin-releasing hormone antagonist (GnRHa) (subcutaneous injection, 240 mg, Degarelix®, Ferring Pharmaceuticals, Sweden) and, after three weeks, to an androgen supplementation by the use of testosterone undecanoate (intramuscular injection, 1000mg, Nebido®, Bayer Pharmaceuticals, Germany) ^20^.

Therefore, blood samples were collected three times: *i*) before TD, corresponding to testosterone and gonadotropins’ baseline levels (Basal group); *ii*) before the androgen supplementation, relating to low testosterone and gonadotropins’ levels (Low group); *iii*) two weeks after TS, corresponding to Testosterone restored samples (Restored group) (Figure 1).

### Sample preparation

Blood plasma samples from the thirty subjects, in fasting condition, submitted to testosterone depletion and replacement were collected in tubes with sodium citrate. For metabolomic experiments, plasma proteins were precipitated with ice-cold methanol enriched with an isotopic labeled internal standard (Testosterone-D3, 50 nM, LGC Standards; London, England). The samples were maintained at -30°C for 30 minutes, followed by centrifugation at 14000g for 15 min at 4°C. Supernatants were collected, dried, and reconstituted in 0.1% formic acid ^27^. A pool of all samples was used as quality control (QC). The QC was prepared using 10 µL of each experimental sample and submitted for the same experimental procedures as performed in the experimental samples. Blank samples (0.1% formic acid) were prepared following the same procedures and used to analyze possible contaminants and to determine background signals. Among the ninety samples, one was excluded from this study due to technical problems.

### UHPLC-HRMS: untargeted metabolomics analysis

Samples were analyzed by ultra-high-performance liquid chromatography (Dionex Ultimate 3000 UHPLC, Thermo Scientific, USA) coupled to high-resolution mass spectrometry (Q-Exactive Plus, Thermo Scientific, USA). Chromatographic separation was carried out employing a C18 column (Zorbax, 50 × 2.1 mm, 1.8 μm, Agilent, USA) and a mobile phase constituted of aqueous solution A (0.1% formic acid and 5mM ammonium formate) and organic solution B (methanol acidified with 0.1% formic acid). The column was held at 40 °C. A gradient method (400 μL.min-1) was applied over 20 min, as follows: 0-1 min, 5% B; 1-14 min, linear gradient from 5 to 100% B; 14-16 min, 100% B; 16-16.5 min, decreasing linearly from 100% to 5% B; 16.5-20 min, 5% B.

The autosampler unit was maintained at 7°C. A volume of 8 µL of each sample was injected in triplicate and analyzed in positive and negative mode. The untargeted metabolomics samples were analyzed in an HRMS with electrospray ionization and scanning in full MS mode (m/z 70-800) with data-dependent acquisition (dd-MS2, top-10 DDA). The source ionization parameters were: spray voltage of 3.90 kV (positive mode) and 2.90 kV (negative mode); capillary temperature, 380 °C; auxiliary gas temperature, 380 °C; 60 sheath gas and 20 auxiliary gas. The mass spectrometer for MS1 and MS2 were set as follows: a) MS1 = resolution of 70 000, AGC target 1e6, maximum IT 100 ms; b) MS2 = resolution of 17 500, AGC target 1e5, maximum IT 50 ms, loop count 10, isolation window 2.0 m/z, HCD normalized collision energy of 30.

Before analyzing the samples, the UHPLC-HRMS system was conditioned with 10 injections of 8 µL of pool plasma, using the same parameters described above. Blank samples and quality controls (QC) were analyzed in the same way as the experimental samples. QC samples were injected and analyzed at six different times during the experiment to assess the equipment’s stability and reproducibility. A washing protocol was applied between each biological sample. Retention time, peak shape, and intensity of the internal standard were monitored to evaluate the system suitability.

### Data processing and metabolomics statistical analysis

Raw files from UHPLC-HRMS were analyzed by Compound Discoverer 3.1 (Thermo Fisher, USA) using an untargeted metabolomics workflow that aligns the MS chromatographic peaks, matches, and compares parents and fragments ions to identify the metabolites present in the samples. The putative metabolites identification was supported by comparison with data from the Human Metabolome Database (HMDB), Kyoto Encyclopedia of Genes and Genomes (KEGG), NIST-Wiley Mass Spectral Library (NIST), Lipid Maps®, BioCyc Database Collection (BioCyc), MassBank, and MZcloud. Thus, the identification of metabolites or putative metabolites was based on a spectrum-structure match of accurate mass and tandem mass data of external libraries, providing a level 2 identification ^52^. Five ppm of mass accuracy, 10^6^ of minimum peak intensity, and 0.2 min of retention time variance were accepted for compound detections.

The average of the triplicate injection was used as a unique value, and the data were normalized by log2 transformation and sample median subtraction. Statistical analyses performed to detect differentially expressed metabolites were carried out in RStudio software. A multilevel sparse partial least squares discriminant analysis (sPLS-DA) was performed using ‘mixOmics’ R package ^53^. This multivariate analysis classifies the samples by performing a multivariate regression while considering the complex structure of repeated measurements. The metabolite abundance matrix (706 plasma compounds) was used as predictors and time point (Basal, Low, and Restored testosterone) as the response variable. The most informative compounds for discriminating samples were selected through a LASSO penalization implemented in the ‘mixOmics’ R package. With the selected compounds, an unsupervised hierarchical clustering plus heatmap was performed using the ‘pheatmap’ R library. In addition to the multivariable analysis, a paired ANOVA (afex::aov_ez R function) followed by FDR correction was performed to determine significant overall differences among time points. To detect differences between individual time points, *i.e*., Basal *vs*. Low, Low *vs*. Restored, and Basal *vs*. Restored, we used the ‘emmeans’ R package for pairwise post hoc multiple comparisons with a *p*-value adjustment equivalent to the Tukey test (obtained through the ‘pair’ R function). Metabolites with an adjusted *p*-value < 0.05 were considered significant. The Log2 fold-change of each comparison was correlated to increment or decrease in metabolites abundance. In this way, negative values were linked to relatively high abundant metabolites and positive values to relatively low abundant metabolites.

All known compounds present in HMDB and KEGG responsible for the cluster separation were employed in the pathways analysis, among them, only six were non-significant (ANOVA paired analysis, *q*-value > 0.05). More information about all compounds pointed by sPLS-DA, such as normalized average area value and Log2 fold-change, is present in the supplementary data (Table S1).

MetaboAnalyst 5.0 was employed to analyze the pathways associated with the statistically significant and deregulated metabolites ^54^. In this sense, metabolic Set Enrichment Analysis based on search in The Small Molecule Pathway Database (SMPDB) library were performed ^55^.

## Data Availability

The data produced in the present study were uploaded to the MetaboLights Platform.

## Funding

The Brazilian National Research Council (CNPq, 308341-2019-8 (G.B.D.); 315167/2020-3 (F.C.S.N)), the Carlos Chagas Filho Rio de Janeiro State Research Foundation (FAPERJ, E-26/210.173/2018 (G.B.D.); 26/202.650/2018 (F.C.S.N.)), and the Interreg V EU program, ReproUnion 2.0 20201846 generously providing funding to support the execution of this study.

## Acknowledgments

We are grateful to the Laboratório de Apoio ao Desenvolvimento Tecnológico (LADETEC) of the Institute of Chemistry from the Federal University of Rio de Janeiro for providing high-quality infrastructure for the UHPLC–HRMS analysis.

## Author contributions

J.S.G., G.M., F.C.S.N., G.B.D. designed metabolomics experiments. J.S.G., G.M. performed metabolomics experiments. J.S.G., I.P., G.M., A.S., F.C.S.N., G.B.D., J.M. analyzed the data. JSG draft the manuscript. K.B.S., I.P., G.M., A.S., F.C.S.N., G.B.D., A.G., R.A., J.M., G.M-V. revised the paper. G.B.D., F.C.S.N., A.G., G.M-V., J.M. funding acquisition. IP and AS performed statistical analyses. J.S.G., I.P., K.B.R., G.M., R.A., G.M-V., A.G., G.B.D., A.S., F.C.S.N., J.M. provided the scientific oversight of the study.

## Data availability

The raw data acquired in this study were uploaded to the MetaboLights Platform.

## Institutional Review Board Statement

This study was registered and approved by the Regional Ethical Review Board Lund (Approval number: DNR 2014/ 311)

## Informed Consent Statement

Informed consent was obtained from all subjects involved in the study

## Conflicts of Interest

The authors declare no competing financial interests

## Notes

### Competing Interest Statement

The authors have declared no competing interest.

### Clinical Trial

NCT03541395

### Funding Statement

Funding: Brazilian National Research Council (CNPq, 308341-2019-8 (G.B.D.); 315167/2020-3 (F.C.S.N)), Carlos Chagas Filho Rio de Janeiro State Research Foundation (FAPERJ, E-26/210.173/2018 (G.B.D.); 26/202.650/2018 (F.C.S.N.)), and the Interreg V EU program, ReproUnion 2.0 20201846 generously providing funding to support the execution of this study.

### Author Declarations

This study was registered and approved by the Regional Ethical Review Board Lund (Approval number: DNR 2014/ 311) and was registered at the NIH clinical trial registry (http://clinicaltrials.gov, Registration number: NCT03541395, date of registration May 17th 2018). Informed consent was obtained from all subjects involved in the study.

