## Supplemental Files: Figures S1-S2, Tables S1-S3 for "Metabolic changes induced by pharmacological castration of young, healthy men: a study of the plasma metabolome": Supplementary Data_Guedes and coauthors_preprint.pdf

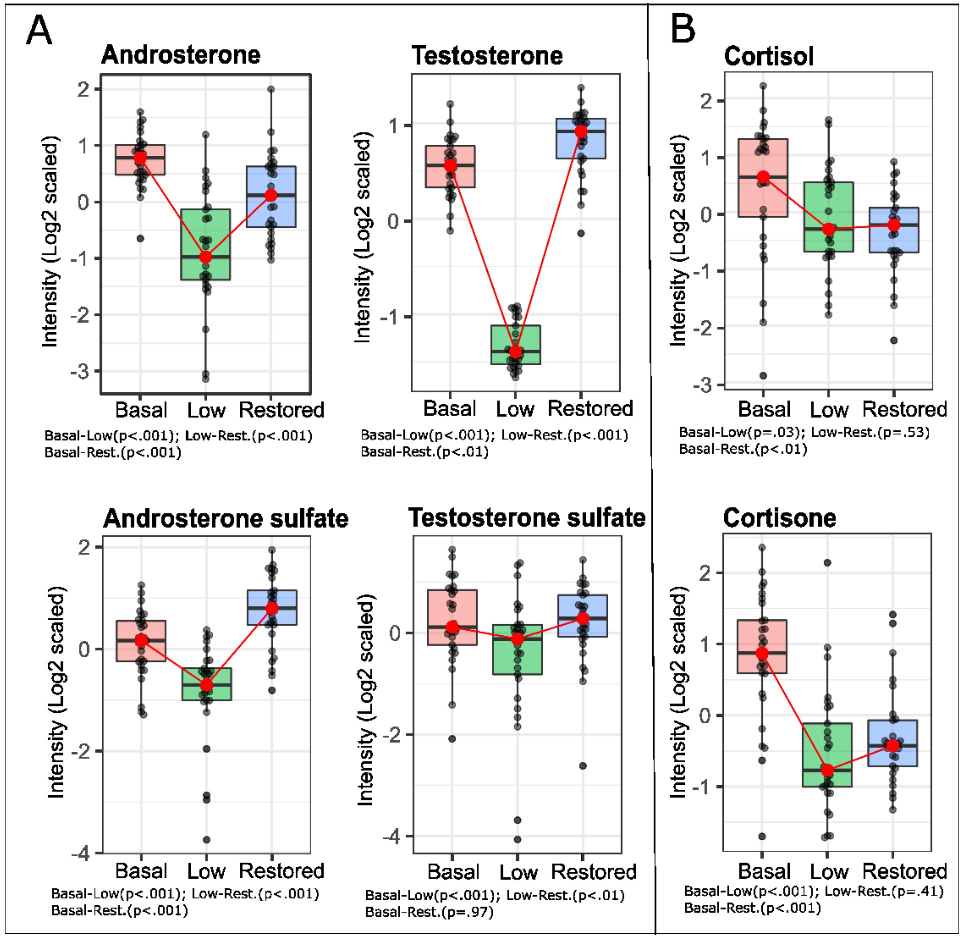


**Figure S1.** Steroids' expression after TD and TS. A) Levels of testosterone and its metabolites. B) Levels of cortisol and cortisone. Boxplot colors: Pink, basal testosterone group; Green, low testosterone group; Blue, restored testosterone group.

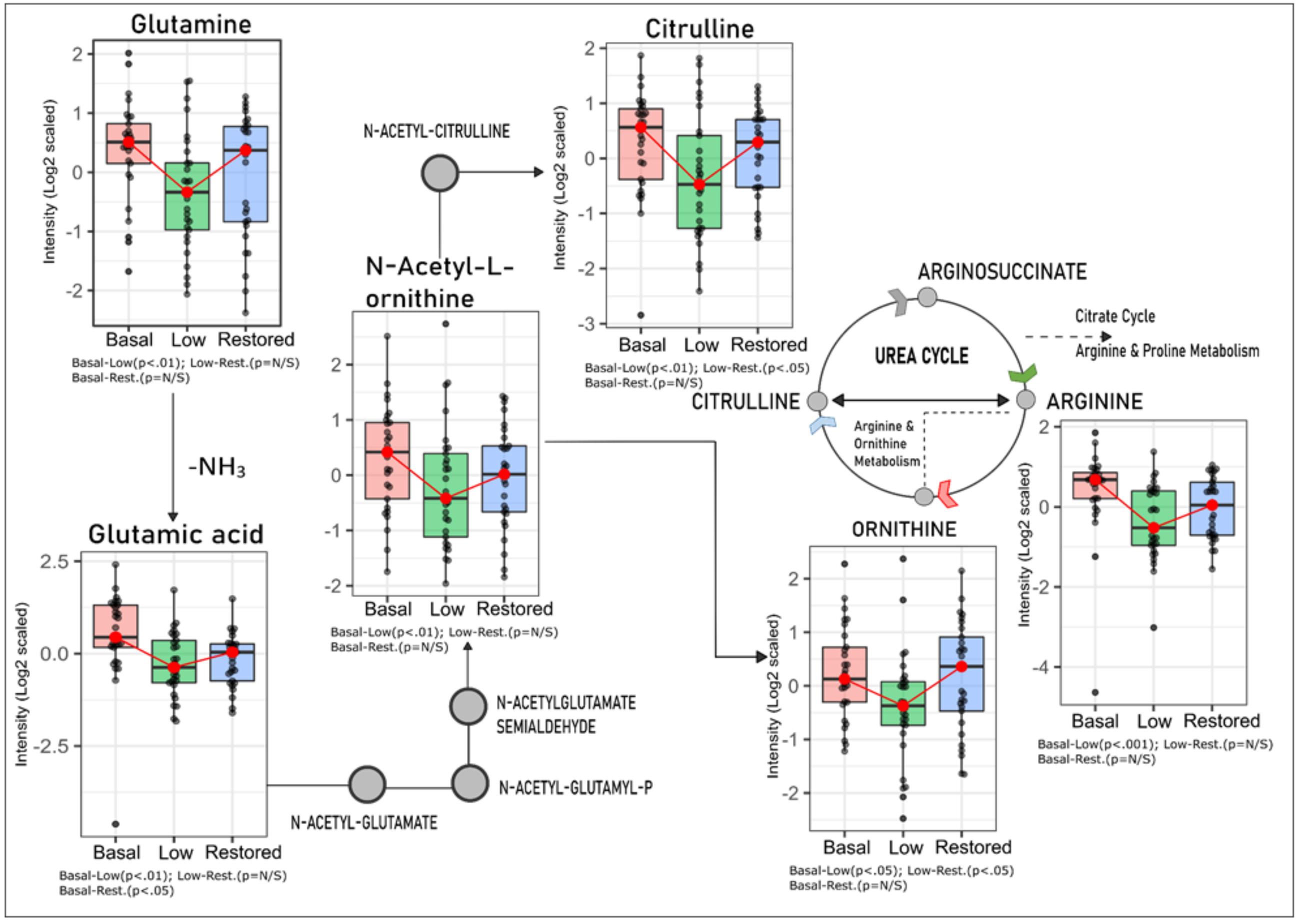


**Figure S2. Testosterone and Arginine Biosynthesis.** Intermediate compounds of arginine biosynthesis present low expression after the ADT. Ornithine and Citrulline were restored after the testosterone replacement ($p$-value <0.05, comparison between Low and Restored group). *N*-Acetylornithine, Glutamine, and Arginine tended to restore based on median values (not statistically significant, comparison between Low and Restored group). Boxplot colors: Pink, basal testosterone group; Green, low testosterone group; Blue, restored testosterone group.
